# Life course circumstances contribute to the acceleration of phenotypic and functional aging in Chinese middle-aged and older adults

**DOI:** 10.1101/2021.09.02.21263060

**Authors:** Xingqi Cao, Chao Ma, Zhoutao Zheng, Liu He, Meng Hao, Xi Chen, Eileen M. Crimmins, Thomas M. Gill, Morgan E. Levine, Zuyun Liu

## Abstract

Accelerated aging implies health inequalities in late life and consequently, poses a huge challenge to society. With two well-validated aging measures, this study aimed to evaluate the relative contributions of life course circumstances to variance in these aging measures with policy implications. We assembled data for 6224 middle-aged and older adults (≥45 years) who participated in the 2014 life course survey, the 2015 biomarker collection, and the 2015 main survey of the China Health and Retirement Longitudinal Study (CHARLS). Two aging measures, including physiological dysregulation (PD) and frailty index (FI), were calculated. Life course circumstances, i.e., 70 circumstances variables involving childhood and adulthood circumstances, demographics, and behaviors, were categorized into 11 study domains for simplicity. The Shapley value decomposition, hierarchical clustering, and general linear regression models were performed. The Shapley value decomposition revealed that all 11 study domains accounted for about 6.3% and 29.7% of the variance in PD and FI, respectively. We then identified six subpopulations who shared similar patterns in terms of childhood and adulthood circumstances. One subpopulation (i.e., disadvantaged) who reported experiencing more childhood and adulthood adversity consistently exhibited accelerated aging indicated by the two aging measures. We conclude that life course circumstances contribute differently to the acceleration of phenotypic and functional aging in Chinese middle-aged and older adults. Special attention should be given to promoting health for the disadvantaged subpopulation and narrowing their health gap with advantaged counterparts. Our findings highlight the role of life course circumstances in ameliorating health inequalities in late life.

## Introduction

Accelerated aging leads to increasing burdens of chronic diseases in late life, posing a huge challenge to society. Geroscience investigators hypothesize that therapies or preventative programs targeting aging would delay the severity or occurrence of most chronic diseases (1). One key step towards success is to measure aging accurately, because aging measures can provide reliable endpoint for which they could be evaluated in preventive and intervention programs of aging. Aging is a multi-dimensional process and has been quantified at hierarchical levels, including biological/molecular, phenotypic, and functional levels (2). Typical examples of summary aging measures include DNA methylation clocks (biological/molecular) (3, 4), Physiological Dysregulation (PD, phenotypic) (5-7), and Frailty Index (FI, functional) (8-10), respectively. These aging measures are different but also share similarities, and might be complementary (11, 12). Overall, these aging measures provide us with an opportunity to evaluate the effect of exposures on the aging process, and further propose potential therapies or preventative programs against aging.

Social-environmental factors, from early life to adulthood, contribute to health inequalities among individuals (13-16), highlighting the importance of adopting a life course approach in aging research. Instead of concentrating on one or a few exposures in certain life stages, a life course approach provides a more complete picture showing how these numerous factors aggregate and synergistically work over a long period. Furthermore, it is well known that childhood and adulthood exposures (e.g., socioeconomic status, SES) may lead to an increased risk of adverse health outcomes in late life through cumulative effects (15, 17). However, the large numbers of life course circumstances pose challenges to statistical analyses that aim to model the effects of individual factors and cumulative effects, and address the high correlation of early life and adulthood factors. In our recent work (13), we have simultaneously assessed the effect of nearly 100 life course circumstances on a measure of phenotypic aging in the US population. We, for the first time, used a series of statistical approaches including the Shapley value decomposition (hereafter, the Shapley method, see details in **S1 Appendix**), hierarchical clustering analysis (HCA), and general linear regression, and successfully addressed the challenges listed above. We demonstrated that these life course circumstances account for about 30% of differences in phenotypic aging.

However, several questions remained unanswered in this work. First, only aging measures at the phenotypic level were considered. Second, only White and Black adults were included and thus, it remains unclear whether the findings apply to other ethnic groups, e.g., the Asians. Third, different nations/countries have their unique social-environmental circumstance, which may change the estimate of effect size. For instance, the Chinese populations are quite different from those in developed countries or other developing countries, in terms of genetics and life course experiences. Many older Chinese have experienced considerable economic and epidemiological transitions (e.g., the Anti-Japan War era, the Great Chinese Famine of 1959-1961, and the reform of the economic system in 1978), which have led to heterogeneity in their aging process.

Facing the rapid population aging in China, we have recently developed a valid physiological biomarkers-based aging measure, PD (at phenotypic level), in the Chinese population (7). PD is robustly predictive of lifespan in the Chinese population and various subpopulations. In addition, FI, a functional aging measure, has been developed in the Chinese population as well (18). FI represents various health and functional domains related to aging, including physical and cognitive function, comorbidity, and symptoms (8). Both PD and FI are significantly associated with an array of health indicators in the Chinese population, including subjective and objective physical functions, and short-term mortality, while the strength differed (**Figure S1**).

With the unique data from the China Health and Retirement Longitudinal Study (CHARLS), for this study, we first assembled over 70 available life course circumstances characterizing childhood and adulthood circumstances, and behaviors in Chinese middle-aged and older adults. Second, we evaluated the relative contributions of these life course circumstances to variance in the two well-validated aging measures. The findings will provide clues for policy programs aimed at alleviating health inequalities and disparities in China, the country with the largest number of older Asians, through targeting multiple exposures across the life course.

## Materials and Methods

### Study population

CHARLS is an ongoing nationally representative and longitudinal survey of Chinese community-dwelling adults aged 45 years and older. CHARLS used a multistage sampling strategy covering 28 provinces, 150 counties/districts, and 450 villages/urban communities across the country. Adults were recruited to the first wave in 2011/2012, and three follow-up waves biennially up to 2017/2018. Details of the CHARLS survey have been described in previous studies (19, 20). In this study, we used data from the 2014 life course survey, the 2015 biomarker collection (20), and the 2015 main survey of CHARLS. As shown in **Figure S2**, 12560 adults participated in all surveys; we excluded those with missing data on each of the biomarkers (N=290) and each of the life course circumstances (N=6046), leaving an analytic sample of 6224 adults. Ethical approval for collecting data on human subjects was received at the Peking University by their institutional review board, and all adults provided informed consent. CHARLS data are publicly available through the CHARLS website (http://charls.pku.edu.cn/en).

### Measures

#### Childhood and adulthood circumstances

In this study, we considered a comprehensive set of factors across the life course (i.e., life course circumstances), which have been associated with adverse health outcomes. The details of questions and responses for these factors are provided in **Table S1**. Briefly, we defined six domains of childhood circumstances (i.e., childhood SES, childhood war, childhood health, childhood trauma, childhood relationship, and childhood parents’ health) and three adulthood circumstances (i.e., adulthood SES, adulthood adversity, and adulthood social support) (**Figure 1**).

**Figure 1.**
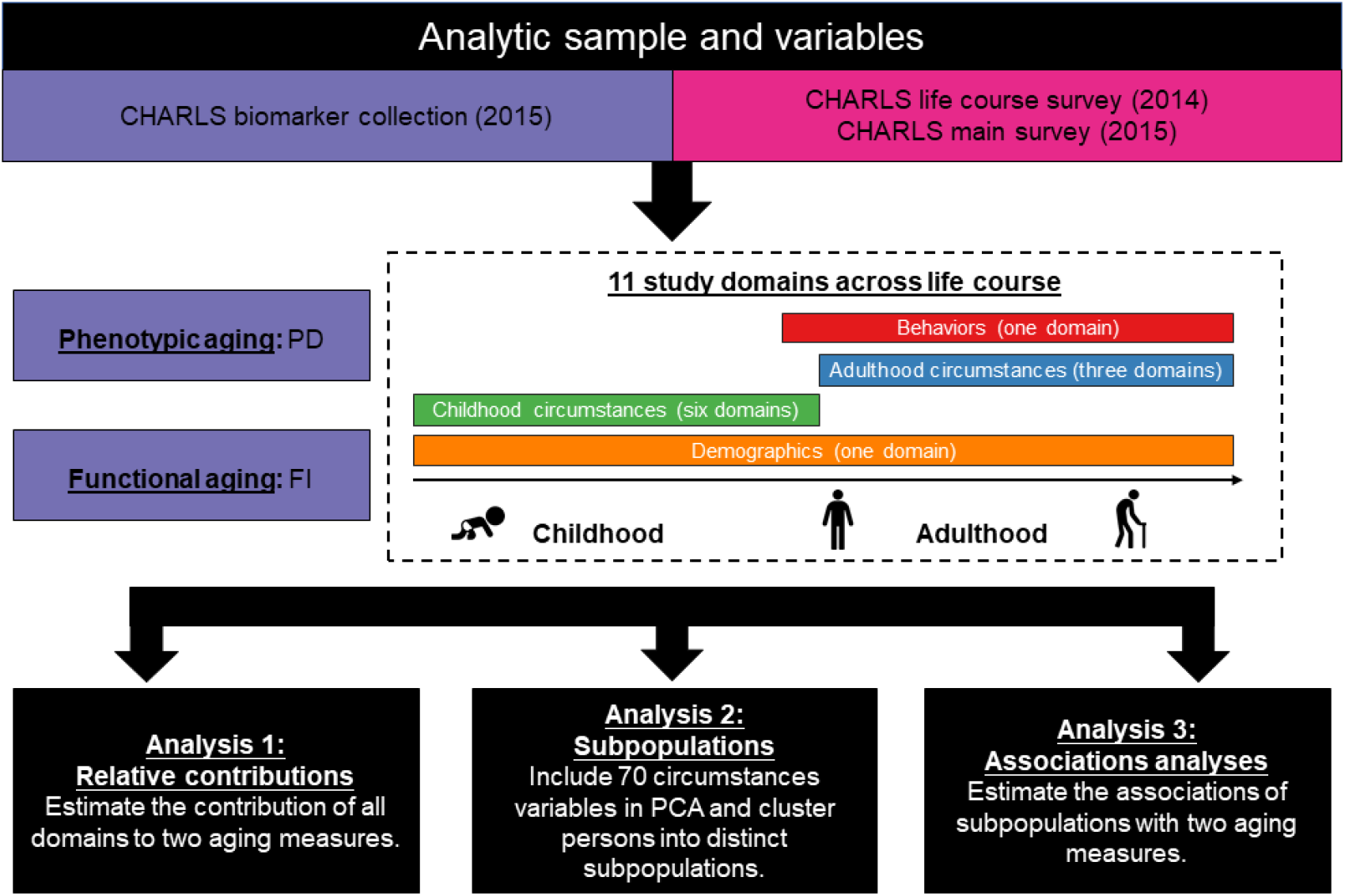
Roadmap for evaluating the association of life course circumstances with two aging measures. The roadmap depicts our analytical procedures. We assembled analytic samples and a large array of variables from the 2014 life course survey, the 2015 biomarker collection, and the 2015 main survey of CHARLS. We categorized a large array of variables across the life course into 11 study domains, including six childhood circumstances domains, three adulthood circumstances domains, one behaviors domain, and one demographic domain. We then performed three analyses to evaluate the association of these domains (particularly the childhood and adulthood circumstances domains) with two aging measures. CHARLS = the China Health and Retirement Longitudinal Study; PD = physiological dysregulation; FI = frailty index; PCA, principal component analysis.

#### Demographics

We considered one domain for demographics, including chronological age, gender, and ethnicity. Ethnicity was classified as Han, and others.

#### Behaviors

We considered one domain for behaviors, including obesity, smoking status, and drinking status. For obesity, we defined a variable called “proportion of obesity”, representing the proportion (range 0-1) of waves at which a respondent had a body mass index (BMI) over 28 kg/m^2^ (13). Smoking status was classified as non-smoker, ever smoker, and current smoker. Drinking status was classified as non-drinker, and drinker.

#### Phenotypic and functional aging measures

We considered two aging measures including PD and FI.

Following the procedure we previously described (7), we calculated PD (derived from Mahalanobis distance). In brief, we used eight biomarkers (i.e., total cholesterol, triglyceride, glycated hemoglobin, urea, creatinine, high-sensitivity C-reactive protein, platelet count, and systolic blood pressure) in the 2015 biomarker collection of CHARLS to calculate Mahalanobis distance, which was then logarithmically transformed (termed as PD in this study) for analysis (5, 6, 21). As done in previous studies, PD was standardized, with no unit. A larger value of PD indicates acceleration of aging and thus a higher risk of mortality (5, 7).

The FI was based on the degree of accumulation of health deficits and represented an alternative measurement of frailty that incorporates health dimensions (e.g., cognition, comorbidities, and disabilities) (8-10). The FI was calculated as the ratio of the number of deficits (variables) out of the total possible deficits (variables) considered (8-10), with a range of 0 to 1. A larger value of FI indicates acceleration of aging. A 39-item version of self-report FI was constructed for CHARLS (18). The list of items included in the FI is provided in **Table S2**. Any adult who had missing data on 20% or less of the variables was retained and the missing values were imputed to the median.

#### Statistical analyses

Characteristics of the total population were presented using mean ±standard deviation (SD) for continuous variables or numbers (percentages) for categorical variables. **Figure 1** briefly summarizes the analytic plan of this study. First, we used the Shapley method to estimate the overall and relative contributions of all life course variables including demographics, childhood and adulthood circumstances, and behaviors to the variance in PD and FI, respectively (see details on Shapley method in **S1 Appendix**).

Second, we used several analyses to cluster adults into distinct subpopulations. We performed principal component analysis (PCA) for 70 circumstances variables. According to the proportional variance explained, we chose the top four principal components and took them as inputs in an HCA, in which we clustered adults into distinct subpopulations. Adults in the same subpopulation shared similar life course circumstances. Then, we compared the two aging measures between different subpopulations to estimate whether accelerated aging occurred in some subpopulations. Finally, we estimated adults’ cluster membership for each subpopulation using a continuous measure that ranged from -1 to 1. To determine the characteristics of each subpopulation, we estimated the correlation between these cluster membership values and circumstances variables.

Third, we used general linear regression models to estimate the associations of identified subpopulations with two aging measures. We documented coefficients and corresponding standard errors (SEs) from two models. Model 1 was unadjusted. Model 2 adjusted for chronological age, gender, proportion of obesity, smoking status, and drinking status. All analyses were performed using SAS version 9.4 (SAS Institute, Cary, NC), R version 3.6.3 (2020-02-29), and STATA version 14.0 software (Stata Corporation, College Station, TX). A P-value <0.05 (2-tailed) was considered to be statistically significant.

## Results

### The characteristics of the study population

The mean chronological age of 6224 adults was 61.6 (SD=8.5) years. About 53% (N=3313) were females, and the majority were Han (93.0%). Details of the characteristics of the study population are presented in **Table S3** and **S1 Appendix**.

### Potential contributions of life course circumstances to phenotypic and functional aging

The Shapley method suggested that the 11 study domains contributed 6.3% (bootstrap SE=-0.0004) and 29.7% (bootstrap SE=-0.006) of the variance in PD (**Figure 2A**) and FI (**Figure 2B**), respectively. All three domains characterizing adulthood circumstances consistently contributed relatively large proportions of variance (PD: 2.7%; FI: 13.1%), with the largest contributor being adulthood SES (PD: 1.5%; FI: 7.1%, rank the first of all 11 study domains, **Figure 2C**). Among the 6 childhood circumstances domains, childhood relationship was the largest contributor (1.1%) for PD, while childhood parent’s health was the largest contributor (2.9%) for FI. Behaviors accounted for 0.1% (rank the tenth), and 2.1% (rank the sixth) of variance in PD, and FI, respectively.

**Figure 2.**
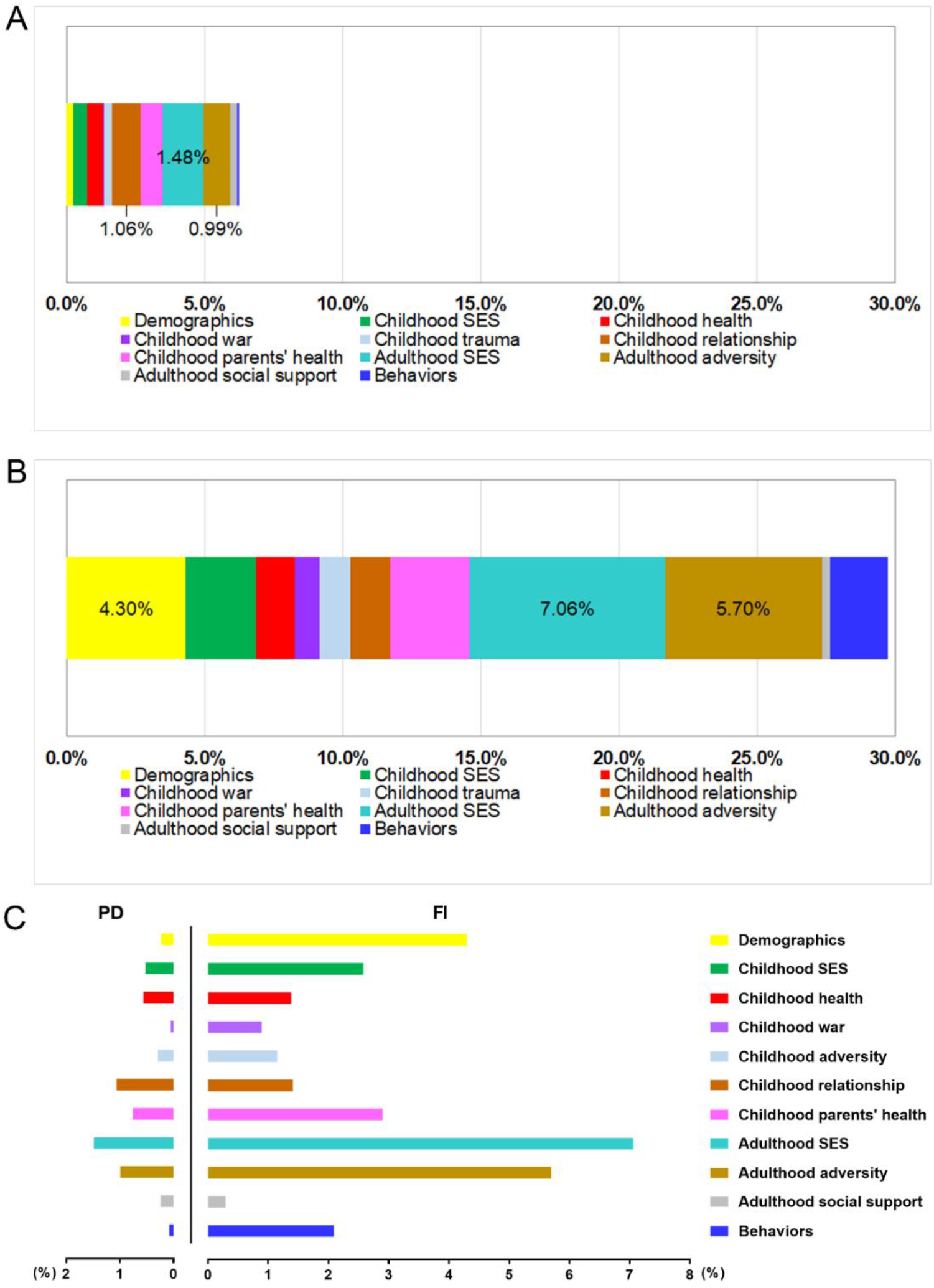
The contribution of all 11 study domains to PD and FI. (A) Stacked bar chart shows the contribution of 11 study domains to PD. (B) Stacked bar chart shows the contribution of 11 study domains to FI. The contribution values of the top three contributors were presented in A and B. (C) Clustered bar chart shows the contribution of 11 study domains to PD and FI simultaneously. The 11 domains include six childhood circumstances, three adulthood circumstances, one behavior, and one demographic. Overall, the 11 study domains contributed 6.3% (bootstrap standard error=-0.0005) and 29.7% (bootstrap standard error=-0.006) of the variance in PD and FI, respectively. SES = socioeconomic status; PD = physiological dysregulation; FI = frailty index.

### Profiles of life course circumstances and subpopulations

Correlations for 70 life course circumstances variables are visualized in **Figure 3A**. We found that childhood SES was mainly correlated with adult SES (evidenced by correlation between education, occupation, and rural residence), but also correlated with factors including being born during a war and childhood neighborhood quality. Childhood trauma was mainly correlated with childhood parents’ health, as well as childhood relationship with friends and parents. We also observed that childhood health was correlated with factors including parents’ depression, childhood relationship with friends and neighborhood, and adulthood adversity.

**Figure 3.**
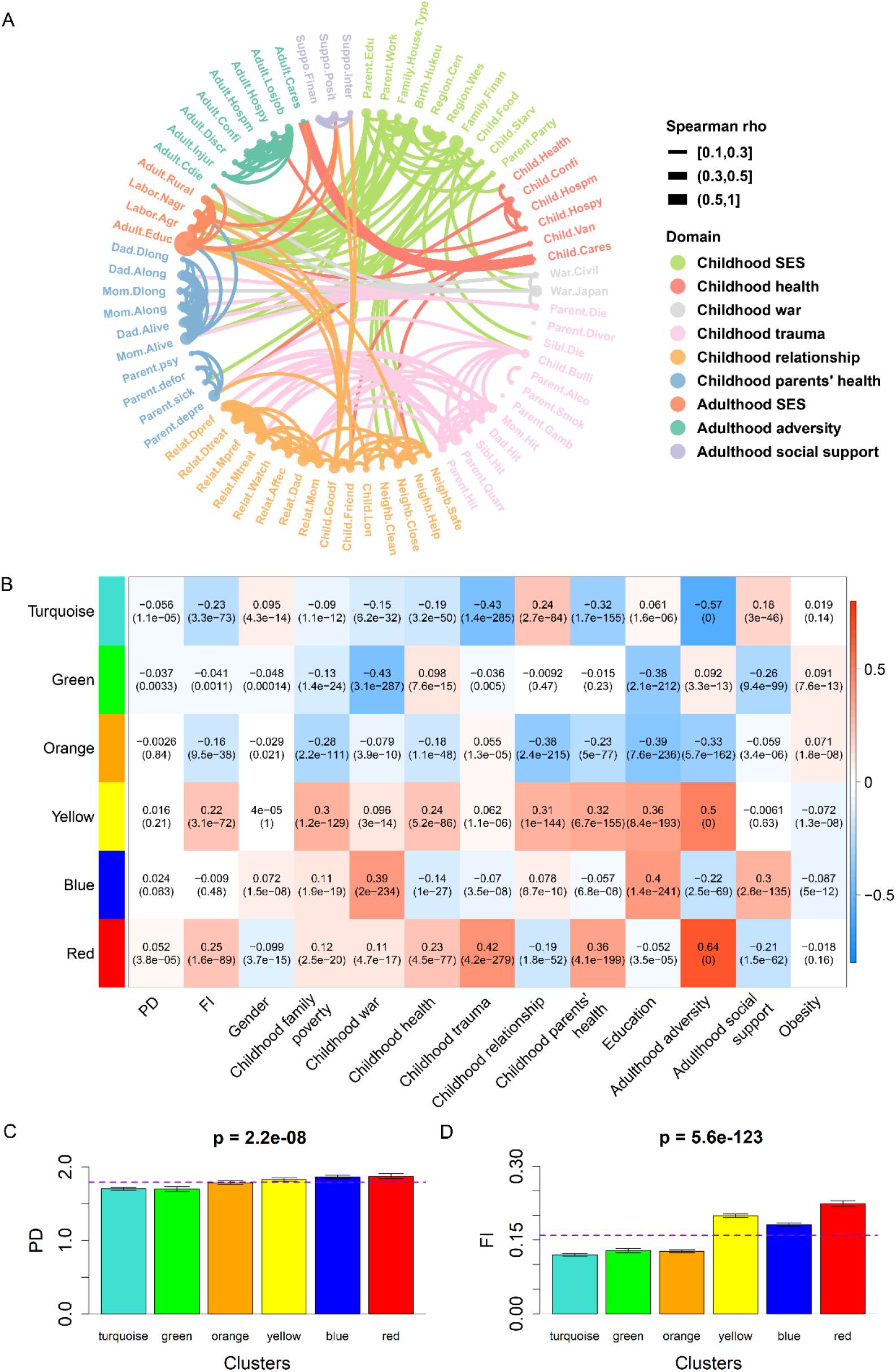
Life course circumstances correlations, cluster membership-trait correlations, and aging measures across six subpopulations. (A) Chord chart of the correlations for each of the life course circumstances. The thickness of ribbon reflects the spearman correlation. (B) Cluster membership-trait correlations and P-values. To determine what each subpopulation/cluster represents, we calculated a continuous measure (cluster membership) for each cluster (between -1 and 1) that denotes how strongly an adult belongs to that given cluster—for example, someone may have a score of 0.9 for the green cluster and-0.7 for the red cluster, suggesting that she/he is very similar to the profile represented by the green cluster, but not the red cluster. Each cell presents the correlation (and P-value) resulting from correlating cluster membership (rows) to traits (columns, including PD, FI, and summarized measures of several circumstances). The table is color-coded by correlation according to the color legend. (C) Bar chart shows PD across the six subpopulations. (D) Bar chart shows FI across the six subpopulations. In C and D, the dotted violet line represents the average level of PD, and FI in the study population, respectively. PD = physiological dysregulation; FI = frailty index.

The correlations of these life course circumstances motivated us to identify six distinct subpopulations from PCA and HCA, each of which was characterized by similar childhood and adulthood circumstances (**Figure S3**). **Figure 3B** shows the profiles of life course circumstances for each subpopulation. Those adults in the red subpopulation were characterized by experiencing more adversity during childhood and adulthood, and having parents with more health problems. The blue subpopulation included adults who were more likely to be born in the war era, had a lower level of education, and acquired less social support during adulthood. The yellow subpopulation included adults who had poor financial situations, had terrible relationship with others, had parents with more health problems during childhood, had a lower level of education, and experienced more adversity during adulthood. In contrast, those in the orange subpopulation were characterized by having a higher level of education, living in great harmony with others, and experiencing less adversity during adulthood. Those in the green subpopulation were not born in the war era, and were more likely to have a higher level of education. Finally, those in the turquoise subpopulation experienced less adversity during childhood and adulthood, and had parents with fewer health problems. The correlations among the six subpopulations are presented in **Figure S4**. We observed an inverse relationship between the turquoise and red subpopulations (r=-0.98), suggesting that the two subpopulations of adults share different life course circumstances. Similarly, the green and blue (r=-0.97), the orange and yellow (r=-0.96) subpopulations were also found to have different life course circumstances.

### Associations of subpopulations with two aging measures

The differences of PD and FI among the six subpopulations are shown in **Figure 3C-D**. The red subpopulation was identified as disadvantaged, with a consistently higher average level of PD and FI. In contrast, the turquoise subpopulation was identified as advantaged relative to the other five subpopulations, indicating that adults in this subpopulation had accelerations of aging.

**Table** 1 presents the formal associations of the six subpopulations with the two aging measures. After adjusting for chronological age, gender, and behaviors, compared with the turquoise subpopulation, the levels of PD and FI in the yellow, blue, and red subpopulations were significantly higher. For instance, relative to the turquoise subpopulation (i.e., advantaged), PD and FI in the red subpopulation (i.e., disadvantaged) were increased by an average of 0.14 (SE=0.04, P<0.001) and 0.10 (SE=0.01, P<0.001), respectively. Obesity was found to be consistently associated with two aging measures, such that proportion of obesity was associated with average increases of 0.07 (SE=0.03) and 0.050 (SE=0.005) in PD and FI, respectively. Compared with non-smokers, FI of ever smokers was increased by an average of 0.03 (SE=0.01, P<0.001).

## Discussion

In a sample of Chinese middle-aged and older adults, we showed that childhood and adulthood circumstances, and behaviors consistently and significantly associated with phenotypic and functional aging, although to a different extent. Overall, the life course circumstances we evaluated accounted for 6.3% and 29.7% of the variance in PD and FI, respectively. Using variables characterizing childhood and adulthood circumstances, we identified six subpopulations, of which the disadvantaged subpopulation exhibiting increased PD and FI. The findings promote the understanding of aging etiology associated with life course circumstances and provide important clues for preventive and interventive programs aiming at slowing the pace of aging through targeting multiple exposures (e.g., those shared by the red subpopulation) across the life course. In addition, the results observed across the two aging measures confirmed that aging measures shared both similarities and differences, suggesting that they may complement each other.

Our results draw attention to several childhood circumstances including being born in the war era, traumatic experience, poor social interrelationship, and parent’s health problems. These childhood circumstances represent the defining features of the disadvantaged subpopulations (i.e., the blue and red subpopulations). While those who had harmonious relationship with others were typical of the advantaged subpopulations (i.e., the orange subpopulation). Prior studies have found that adults who live in hostile neighborhoods have relatively shorter telomere length, which is associated with accelerated cellular aging (22, 23). Childhood negative events may have become biologically embedded in the disruption of functioning across body systems, including immunity, inflammation, and metabolism (24). In both human and animal studies, early life adversity has been linked to immune dysregulation over the life course (25-28). Adults who experienced adversity in childhood have enhanced stress sensitivity and fewer available psychological and social resources, thereby exhibiting greater physiological deterioration in the immune system (25). Our findings suggest that exposures to traumatic experiences and parent’s health problems in childhood especially contribute to accelerated phenotypic and functional aging. Hence, preventing or reducing the severity of these negative early-life events may help reshape the trajectory of the population towards healthier aging.

Childhood SES (e.g., childhood family poverty) served as an important contributor to aging in this study, consistent with previous studies (29, 30). The poor economic condition in childhood could induce biological damages that increase susceptibility to disease (31-33). Low SES could trigger high levels of stress hormones (e.g., catecholamines, and cortisol) (32), which may accelerate the aging process (33). Also, childhood SES may be linked to accelerated aging through exposure to malnutrition, infection disease, and unavailability of medical care. Our results highlight the need for special attention to children from low SES families to diminish health inequality in late life.

As with childhood SES, numerous studies have focused on the influences of adulthood SES and adversity on aging (34-36). However, a recent study found that the positive influence of childhood SES on late life health trajectory was indirectly established through its influence on adulthood SES (37). Adults with poor childhood economic conditions may subsequently experience more adversities in adulthood. Thus, the associations of circumstances in childhood and adulthood with aging are difficult to disentangle with traditional statistical methods. In this study, we were able to assess the relative contribution of life course circumstances using the Shapley method, so as to provide a relatively accurate estimate for the above associations. We found that both childhood and adulthood circumstances appear to contribute to variance in aging, suggesting that we should keep a watchful eye on circumstances over the life course, and highlighting the importance of life course preventative programs and policy.

Consistent with an earlier study in the US population (13), we demonstrated that behaviors contribute to accelerated aging (**Table 1** and **Figure 2**), highlighting the importance of adherence to healthy behaviors to promote healthy aging. Particularly, BMI, as an indicator of obesity, has been found to be positively associated with faster epigenetic aging as measured by DNA methylation (38, 39). This is similar to our observations of proportion of obesity in this study. Given that a recent intervention trial found that lifestyle weight loss may attenuate DNA methylation age (40), it is highly recommended to propose policy programs targeting behaviors (modifiable factors) to improve population health.

**Table 1.**
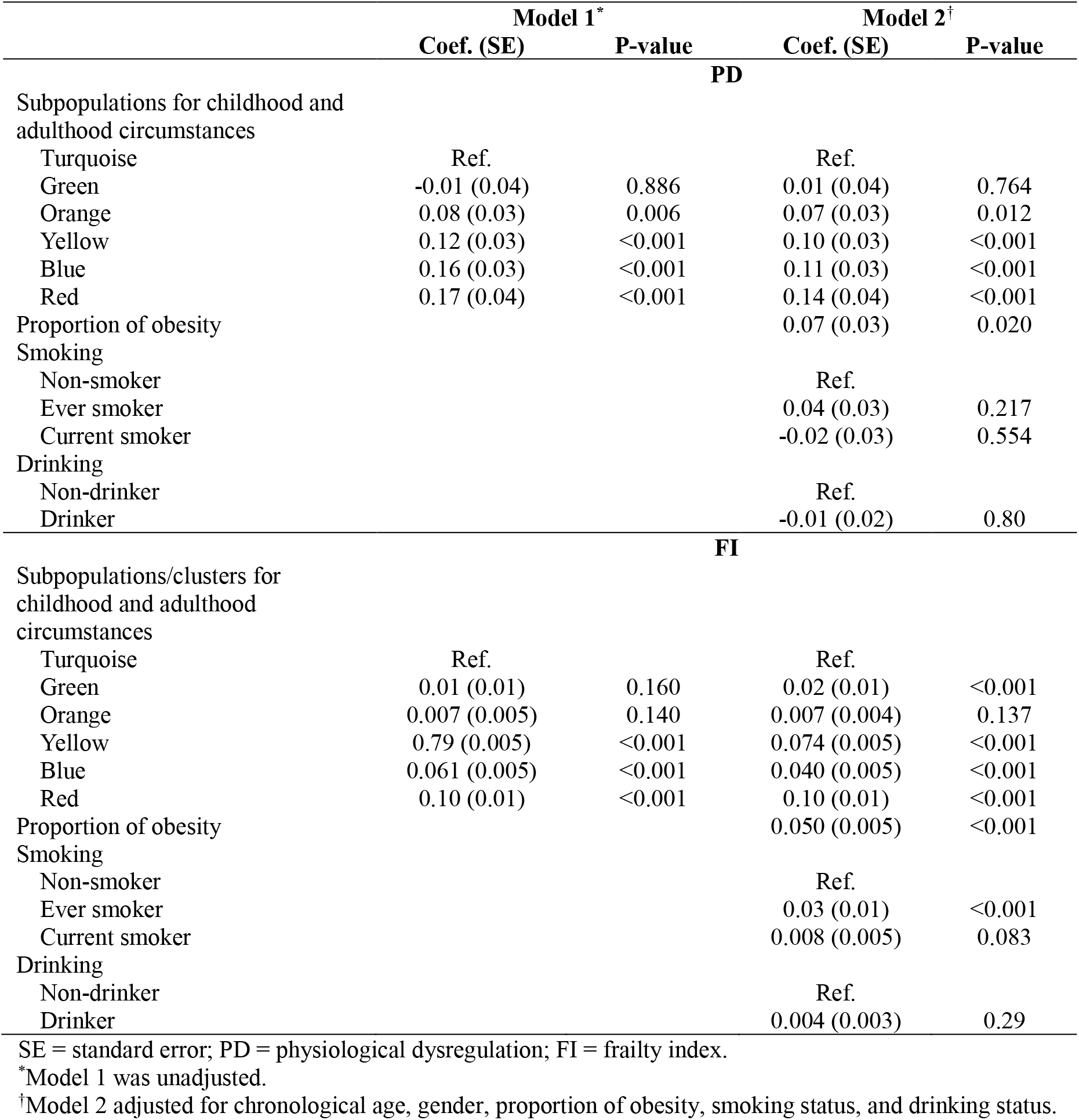
Associations of childhood and adulthood circumstances with two aging measures.

|  | Model 1* |  | Model 2† |  |
| --- | --- | --- | --- | --- |
|  | Coef. (SE) | P-value | Coef. (SE) | P-value |
| **PD** |  |  |  |  |
| Subpopulations for childhood and adulthood circumstances |  |  |  |  |
| Turquoise | Ref. |  | Ref. |  |
| Green | -0.01 (0.04) | 0.886 | 0.01 (0.04) | 0.764 |
| Orange | 0.08 (0.03) | 0.006 | 0.07 (0.03) | 0.012 |
| Yellow | 0.12 (0.03) | <0.001 | 0.10 (0.03) | <0.001 |
| Blue | 0.16 (0.03) | <0.001 | 0.11 (0.03) | <0.001 |
| Red | 0.17 (0.04) | <0.001 | 0.14 (0.04) | <0.001 |
| Proportion of obesity |  |  | 0.07 (0.03) | 0.020 |
| Smoking |  |  |  |  |
| Non-smoker |  |  | Ref. |  |
| Ever smoker |  |  | 0.04 (0.03) | 0.217 |
| Current smoker |  |  | -0.02 (0.03) | 0.554 |
| Drinking |  |  |  |  |
| Non-drinker |  |  | Ref. |  |
| Drinker |  |  | -0.01 (0.02) | 0.80 |
| **FI** |  |  |  |  |
| Subpopulations/clusters for childhood and adulthood circumstances |  |  |  |  |
| Turquoise | Ref. |  | Ref. |  |
| Green | 0.01 (0.01) | 0.160 | 0.02 (0.01) | <0.001 |
| Orange | 0.007 (0.005) | 0.140 | 0.007 (0.004) | 0.137 |
| Yellow | 0.79 (0.005) | <0.001 | 0.074 (0.005) | <0.001 |
| Blue | 0.061 (0.005) | <0.001 | 0.040 (0.005) | <0.001 |
| Red | 0.10 (0.01) | <0.001 | 0.10 (0.01) | <0.001 |
| Proportion of obesity |  |  | 0.050 (0.005) | <0.001 |
| Smoking |  |  |  |  |
| Non-smoker |  |  | Ref. |  |
| Ever smoker |  |  | 0.03 (0.01) | <0.001 |
| Current smoker |  |  | 0.008 (0.005) | 0.083 |
| Drinking |  |  |  |  |
| Non-drinker |  |  | Ref. |  |
| Drinker |  |  | 0.004 (0.003) | 0.29 |

SE = standard error; PD = physiological dysregulation; FI = frailty index.
\*Model 1 was unadjusted.
†Model 2 adjusted for chronological age, gender, proportion of obesity, smoking status, and drinking status.

Although our results consistently demonstrated the impact of life course circumstances on aging, we should also note the differences. First, the total contribution of these factors varies greatly with respect to the different aging measures (29.7% for FI, but only 6.3% for PD). Second, the same factor differentially contributes to the variance for each of the two aging measures. For instance, the behaviors ranked the sixth in 11 contributors for FI, while it was the penultimate contributor for PD. These differences could be explained by the heterogeneous information that aging measures covered (multisystem clinical biomarkers vs. function aspects) (41) and algorithms used to develop aging measures (e.g., Mahalanobis distance vs cumulative approach). Our findings suggest that these aging measures share both differences and similarities (11, 12, 41). In moving forward, broader research should leverage the potential of multiple aging measures to explore the reasons and consequences of the aging process.

On the basis of our previous study in the US population, this study contributes to the literature in several aspects. First, we used two aging measures, resulting in novel findings as mentioned above. Second, we showed that the observations in Whites and Blacks did not appreciably apply to this group of Asians. This was evidenced by the different magnitude of contributions of factors (e.g., behaviors) to phenotypic aging in the US and China. Third, we draw attention to many unique factors such as being born in the war era, rural residence, those only the Chinese have and the policy could target.

A major strength of this study is the availability of data from a large nationally representative sample of the middle-aged and older Chinese, providing a unique opportunity to estimate the contributions of a comprehensive set of life course circumstances to aging in China. The world’s largest aging population in China has special features, such as having experienced considerable economic and epidemiological transition during their lives. Therefore, our results offer important evidence for life course management to promote healthy aging in China. The second strength of the study is the novel approaches (e.g., the Shapley method) we used to address challenges raised by large numbers of life course circumstances. Finally, we simultaneously considered two aging measures in the same population and examined their associations with life course circumstances, which has not been done in prior research.

This study also has limitations. First, many circumstances variables were based on self-reports, and thus, our results may be influenced by recall bias. Second, the study included Chinese middle-aged and older adults, those born in that period with high mortality, which may introduce survival bias. Third, we did not have the specific timing of childhood and adulthood circumstances, which may affect the effect of circumstances on aging. Fourth, we were unable to cover all life course circumstances variables and confounding variables, leading to possible underestimations of the overall contribution to aging. Fifth, the cross-sectional measurements of aging impeded us to track differences in rates of aging, which could be addressed in the future with longitudinal data. Finally, a large proportion of adults were excluded due to missingness, partially affecting the representativeness of the study population (**S1 Appendix**).

In summary, despite the varying contributions, life course circumstances were associated with accelerations of phenotypic and functional aging in Chinese middle-aged and older adults. We estimated that demographics, behavioral, social-environmental circumstances over the life course account for about 6%-30% of the variance in aging. Furthermore, we identified a vulnerable subpopulation who experienced childhood and adulthood adversity and exhibited accelerated aging. The findings highlight the role of life course management in slowing aging and thus in ameliorating health inequalities in late life. In moving forward, studies should leverage aggregated and concurrent life course circumstances to develop potential therapies or preventative programs of aging. Meanwhile, special attention should be given to promoting health for the disadvantaged subpopulation and narrowing their health gap with advantaged counterparts.

## Data Availability

The the China Health and Retirement Longitudinal
Study (CHARLS) data are publicly available through the CHARLS website at
http://charls.pku.edu.cn/en.

http://charls.pku.edu.cn/en

## Author Contributions

Z.L. conceived and designed research; X.C., C.M., Z.Z., L.H., M.H., Xi Chen, E.M.C, T.M.G, M.E.L, and Z.L. performed research; X.C., C.M., Z.Z., L.H., M.H., and Z.L. analyzed data; and X.C., C.M., Xi Chen, E.M.C, T.M.G, M.E.L, and Z.L. wrote the paper. All authors have read and agreed to the final version of the manuscript.

## Data Availability Statement

The the China Health and Retirement Longitudinal Study (CHARLS) data are publicly available through the CHARLS website at http://charls.pku.edu.cn/en.

## Acknowledgments

We appreciate all adults who participated in the China Health and Retirement Longitudinal Study.

